# The in-frame p.Lys254del-CAPN3 deletion is not sufficient to cause late-onset camptocormia in dominantly inherited calpainopathy

**DOI:** 10.1101/2023.07.19.23292361

**Authors:** Andrea Valls, Gerardo Gutiérrez-Gutiérrez, Agustín Martínez, Cristina Ruiz-Roldán, Pilar Camaño, Adolfo López de Munain, Amets Sáenz

## Abstract

**Introduction/Aims:** Limb-girdle muscular dystrophy R1 (LGMDR1) calpain 3-related is one of the most common forms of LGMD. It is typically recessively inherited and associated with progressive weakness of proximal limb-girdle muscles. Recently, several families with an autosomal dominant inheritance transmission pattern have been reported (LGMDD4). Camptocormia is a common clinical feature in these patients. In these families, different mutations in *CAPN3* have been identified, including in-frame deletions and missense mutations. In particular, two patients presenting with camptocormia carried the p.Lys254del missense mutation without a second identified mutation in *CAPN3*.

**Methods:** Among our patients, we identified two LGMDR1 families as carriers of the p.Lys254del mutation by DNA sequencing, both in homozygous and compound heterozygous states and clinically analyzed the family members carrying this mutation.

**Results:** Interestingly, these patients did not present the myopathic characteristics described in the so-called dominant patients. No camptocormia or any other severe clinical symptoms were observed.

**Discussion:** Accordingly, we conclude that the p.Lys254del mutation *per se* cannot be solely responsible for the camptocormia observed in dominant patients. Other additional undisclosed factors might regulate the phenotype associated to a dominant inheritance pattern in *CAPN3* mutation carriers.

## Introduction

Recessively inherited limb-girdle muscular dystrophy R1 (LGMDRI) calpain 3-related, one of the most common forms of LGMD, is typically associated with progressive weakness of proximal limb-girdle muscles ^1,2^. LGMDR1 is secondary to mutations in *CAPN3* ^3^, the gene that encodes calpain-3, which is expressed predominantly in skeletal muscle ^4^.

In 2012, Liewluck et al. ^5^ had already described a case with axial myopathy and camptocormia bearing a mutation in *CAPN3*. Vissing and colleagues ^6^ were the first to identify families with an autosomal dominant inheritance pattern, with a single mutation in *CAPN3*; however, since then, numerous works have described families showing this transmission pattern ^7–13^.

Although the reason for the different transmission pattern is still unknown, some authors propose that the only found mutation in *CAPN3* gene may have a dominant negative effect, not allowing normal proteins to carry out their function due to the presence of mutated versions of the protein in the muscle fiber ^6,10^.

Two studies reporting on this inheritance pattern described two patients bearing the p.Lys254del mutation without a second identified mutation in *CAPN3* who showed camptocormia ^5,13^. Camptocormia is defined as a pathological involuntary flexion of the thoracic and lumbar spine. It is commonly caused by a wide range of neurological diseases, particularly movement and neuromuscular disorders ^14–17^.

In this study, we report on two LGMDR1 families including carriers of the p.Lys254del mutation, one in homozygosity and the other in a compound heterozygous state. Contrary to previous reports, neither the carriers of this mutation, nor the parents or siblings of the propositus cases in our families present any clinical or radiological characteristics previously associated to the so-called dominant patients. Accordingly, bearing the p.Lys254del mutation alone cannot explain the camptocormia observed in dominant patients; thus, other unveiled genetic factors probably determine the phenotype and the transmission pattern in these carriers.

## Materials and methods

LGMDR1 patients carrying the p.Lys254del mutation as well as their families were recruited from the Hospital Universitario Infanta Sofía (Madrid) and Hospital Público da Mariña - Burela (Lugo). The study was approved by the Comisión de Investigación del Hospital Universitario Infanta Sofía (Madrid) and Comité de Ética del Área Sanitaria de Lugo, A Mariña y Monforte (Lugo); ethical approval was given and all subjects provided written informed consent.

### Family A

The patients were two sisters, homozygous for the p.Lys254del mutation, born of consanguineous parents. They showed the typical symptoms of LGMDR1. First symptoms were reported during the second decade of life, when they started having problems keeping up with their peers. The muscle weakness started proximally and progressed swiftly. In the late adolescence, they could not climb stairs and by their 20’s needed a wheelchair. The patients presented a classic calpainopathy phenotype with higher proximal than distal weakness in both upper and lower limbs, scapular winging, lumbar hyper lordosis, and bilateral ankle dorsiflexion weakness. Currently, in their late 30’s and 40’s, they both use an electric wheelchair for most of the day.

### Family B

In this family, the two affected cases were compound heterozygous for p.Lys254del/ p.*822Leuext*62 mutations. The age at onset for the oldest brother, patient II-2, was in his first decade of life. He showed difficulties to stand up from a sitting position and exercise, and reported frequent falls. Ever since, he showed a slow progression. At the time of the study (late 30’s), he had severe weakness of the pelvic and scapular girdles as well as marked bilateral Achilles tendon contractures. His brother, patient II-4, presented symptoms first in his second decade of life. Two years later, he showed difficulty to stand up from a sitting position. His progression is slow as well but he presented a less severe phenotype than his brother. No other family members were affected.

After obtaining the patient informed consent, DNA/RNA were extracted from peripheral blood samples according to standard procedures. Amplification of *CAPN3* coding regions was performed as in Richard and colleagues ^3^. For the cDNA analysis, we used a previously described protocol ^18^.

## Results

From our series of patients with LGMDR1, nine cases in six families carried the p.Lys254del mutation in a homozygous or compound heterozygous state. However, only two out of the six families were available for the study of mutation carriers.

### Family A

In this family, the two affected patients were homozygous for the p.Lys254del mutation. To establish whether the relatives were also carriers, the DNA of the consanguineous parents and the other sibling was analyzed. The carrier status was confirmed in all of them (Fig. 1 and Table 1). In addition, a possible muscle impairment was studied in the three carriers (Fig. 2). The father, in his 70’s did not show any muscular symptoms. However, he presented an elevated Creatine Kinase (CK) (981U/L) (NV: 46–171 U/L), although this could be partially due to a moderate Obstructive sleep apnea-hypopnea syndrome (OSAHS). He had a normal spirometry, and magnetic resonance imaging (MRI) of the thighs and paraspinal muscles showed no abnormal findings (Fig. 2B). The mother, in her 70’s showed a normal CK level (85 U/L) and the MRI findings of the thighs were normal (Fig. 2C) (an MRI of the paraspinal muscles was not available); however, she suffered from pseudofibromyalgia. The brother in his 40’s carried the mutation and had slightly elevated CK levels (254 U/L). However, no muscular symptoms were observed and the MRI findings of the thighs were normal (Fig. 2C).

**Fig. 1:**
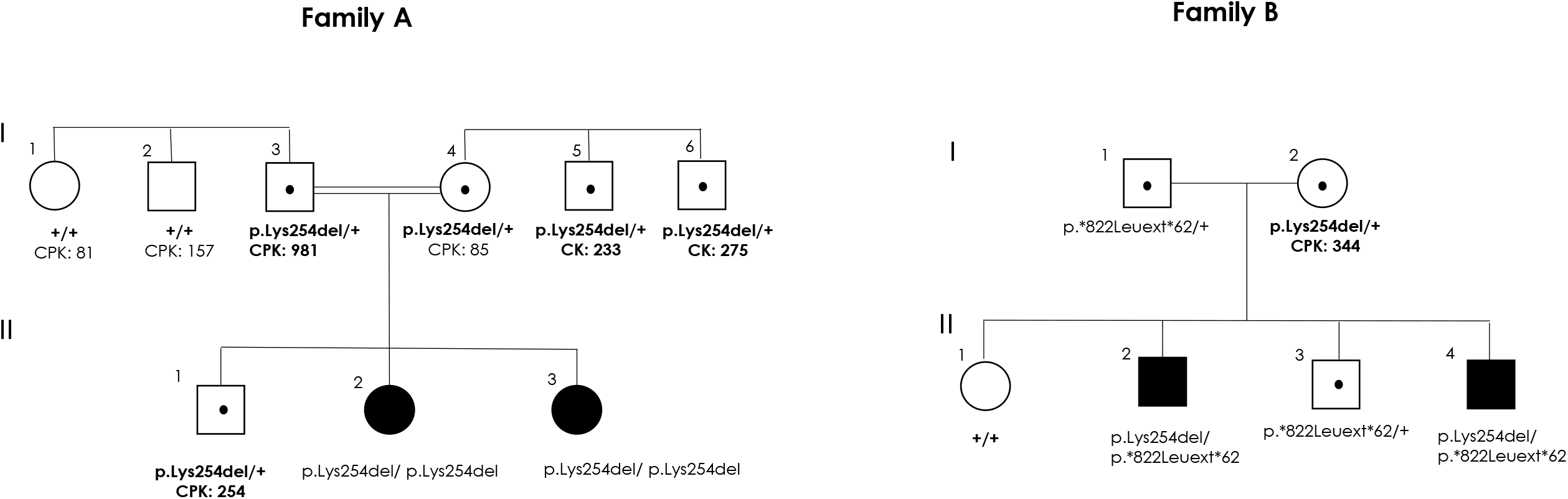
Pedigrees of the two families carrying the p.Lys254del mutation. Filled symbols designate LGMDR1 patients. Open symbols with a black dot designate p.Lys254del mutation carriers.

**Table 1:** p.Lys254Δ mutation carriers’ clinical information. CPK Normal value : <171 U/L. N.A: Not available.

| Family A | Mutation in *CAPN3* gene | Age | CPK | Clinical symptoms | MRI |
| --- | --- | --- | --- | --- | --- |
| I-3 | p.Lys254Δ/+ | 76-80 | 981 | Moderate OSAHS<br>Normal spirometry | Normal thighs and paraspinal muscles |
| I-4 | p.Lys254Δ/+ | 66-70 | 85 | Pseudofibromialgia<br>No muscular dystrophy | Normal thighs |
| I-5 | p.Lys254Δ/+ | 76-80 | 233 | No muscular dystrophy | N.A. |
| I-6 | p.Lys254Δ/+ | 66-70 | 275 | No muscular dystrophy | N.A. |
| II-1 | p.Lys254Δ/+ | 41-45 | 254 | No muscular dystrophy | Normal thighs |
| **Family B** |  |  |  |  |  |
| I-2 | p.Lys254Δ/+ | 66-70 | 344 | No muscular dystrophy | N.A. |

**Fig. 2:**
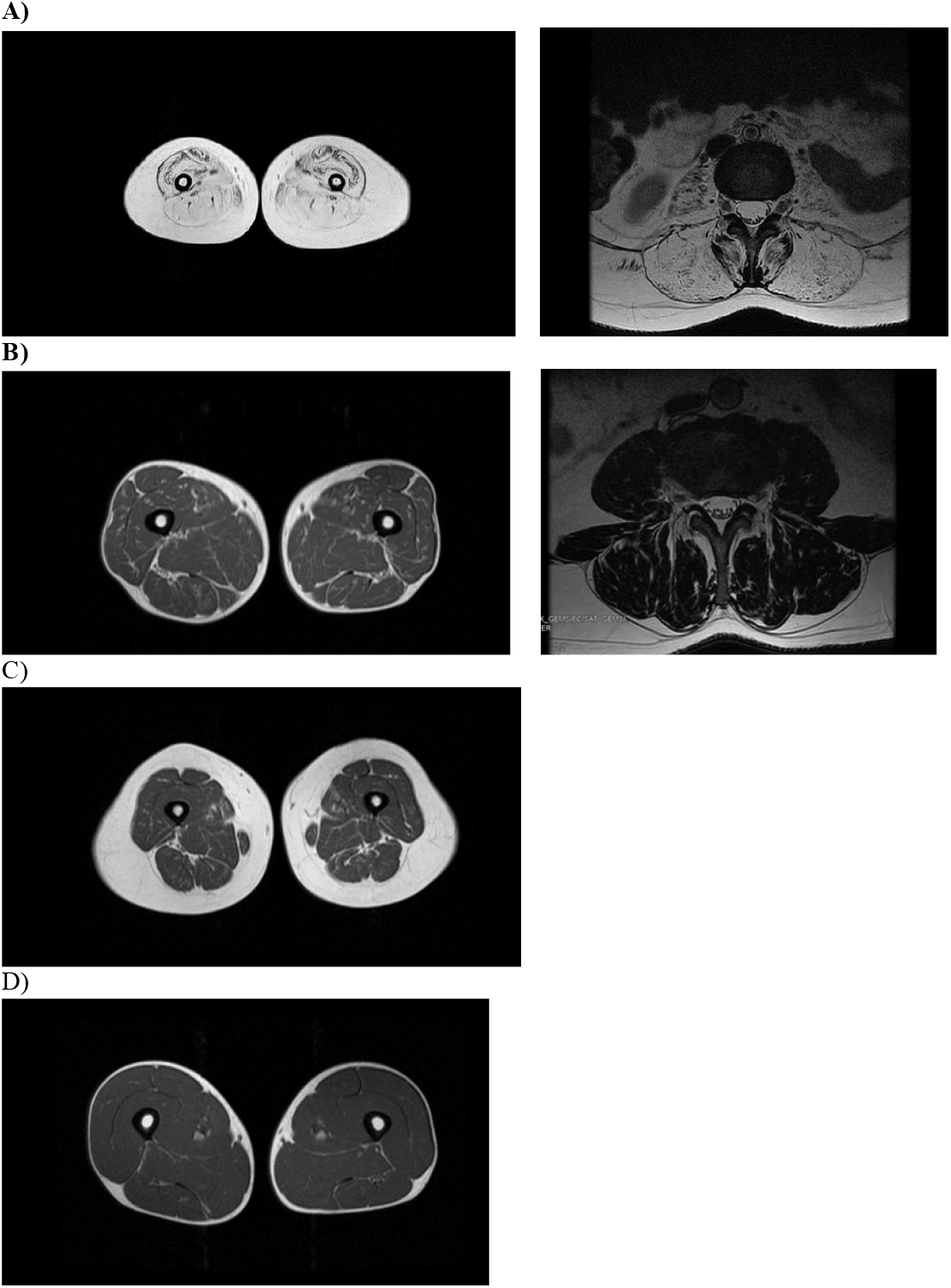
MRI of Family A members. (**A)** II-2 patient (Homozigous for the p.Lys254del mutation): thighs and paraspinal muscles. (**B)** I-3 carrier father, thighs and paraspinal muscles. (**C)** I-4 carrier mother, thighs. (**D)** II-1 carrier brother, thighs.

In addition, other family members, that is, the parents’ siblings were analyzed to establish their carrier status (Fig. 1). Two of the brothers from the mother (I-5 and I-6) were carriers of the p.Lys254del mutation and presented moderately increased CK levels, 233 and 275 U/L, respectively.

### Family B

Healthy family members were analyzed to characterize their carrier status. The oldest sibling did not carry any mutation but the father and the other sibling were carriers of the p.*822Leuext*62 mutation (Fig. 1). In this family, the mother was the only carrier of the p.Lys254del mutation; she showed a CK level of 344 U/L, but did not show any myopathic symptoms (Table 1).

Finally, in our calpainopathy patients’ series, we found two *CAPN3* gene mutation carriers with increased CK levels but no muscular symptoms. One is a patient’s mother (CK: 244), carrier of the p.Gly222Arg mutation and the other is a patient’s brother (CK: 510), carrying the p.Arg788Serfs*14 mutation.

## Discussion

The fact that a large number of families with an autosomal dominant inheritance pattern has been described ^5–7,9–13,19^ confirms the existence of this new calpain 3 related muscular dystrophy form. In other muscular dystrophies, the possibility of a dominant or recessive inheritance pattern associated with mutations in the same gene has already been described, as in titinopathies or in RYR1-related myopathies ^20–24^. However, the mechanisms underlying these different inheritance patterns remain unclear for many of them.

Examination of the increasing number of cases with only one mutation in *CAPN3* gene might help clinicians better understand the dominant phenotype, which differs from the typical LGMDR1 phenotype. This phenotypic difference might indicate the involvement of an additional modulating factor, as a second uncharacterized mutation in *CAPN3* seems an unlikely possibility.

*CAPN3* mutation carriers (healthy relatives of recessive forms bearing only one mutation) that show no muscle atrophy or any severe clinical signs, suggest that the muscle function is not compromised. That is, the amount of protein produced by the healthy allele allows maintaining muscle fiber function. However, could the muscle still maintain its correct activity with an additional mutation in another gene encoding a protein involved in any other muscle function? Previously, this seemed like a *non sequitur*, as various genes causing known muscular dystrophies were ruled out ^7,9–12,19^. However, the intronic sequences of these genes were not considered, nor have been other genes without clear involvement in muscular dystrophies but encoding proteins with diverse cellular functions, such as metabolic functions, signal transduction, channels, etc.

The requirement of a regulatory or modifying factor appears supported by the observation that our mutation carriers do not present muscle impairment. Most carrier members of the two families analyzed are beyond the age of onset of the common symptomatology described for dominant forms. Finally, the normal MRI of the paraspinal muscles demonstrated that camptocormia is not a direct consequence of the presence of p.Lys254del mutation.

Moreover, the two p.Lys254del mutation carriers previously reported presented phenotypic differences ^5,13^. These patients presented paravertebral involvement, however, even though these patients showed similar age and years of evolution, they showed differences in thigh muscle impairment. In the patient presented by Liewluck and colleagues ^5^, there was involvement of the gluteus and the anterior compartment of the thighs, but not in the patient described by Spinazzi and colleagues ^13^. In our studied family members, there was no impairment of the thighs. This suggests that additional regulatory or modifying factors are involved in muscle function impairment, which could potentially explain the phenotypic differences between the cases introduced by Liewluck et al. ^5^ and Spinazzi et al. ^13^.

In recessive muscular dystrophies, such as α-sarcoglycanopathies and dysferlinopathies, carriers can present a subclinical symptomatology (elevated CK levels and abnormal MRI findings) ^25–27^. All our p.Lys254del carrier members, showed elevated CK levels except for the case I-4 in Family A. This suggests that the increased CK level is not only due to the p.Lys254del mutation but additional factors may be responsible for the increased CK levels. In our calpainopathy patients’ series, we found two *CAPN3* gene mutation carriers with increased CK levels but no muscular symptoms. One of the cases carried the p.Arg788Serfs*14 mutation. Since the p.Arg788Serfs*14 mutation results in a total absence of CAPN3, we consider that it could not cause a dominant negative effect (increased CK level) in healthy carriers.

Previous studies suggest that the nature of the mutation is the most plausible reason for dominant transmission in calpain 3-associated dystrophies ^6,10^. However, to confirm that this is the case more in-depth research is needed given the increasing number and type of mutations associated with this form. As examples, cell-based proteolytic assays indicated a loss of autolytic and proteolytic function in p.Lys254del mutants but a dominant negative functional effect was eliminated ^13^. Similarly, the p.Arg572Pro mutation described by Vissing et al. ^12^ is not responsible for causing a dominant negative effect. The *CAPN3* wild type co-transfection assay demonstrated that the mutation does not cause the loss of proteolytic activity of the wild type form. Based on these findings, Spinazzi et al. ^13^ considered that the molecular mechanism underlying dominant forms is likely related to haploinsufficiency, which may constitute a risk factor for paravertebral myopathy with aging.

In conclusion, our results strongly suggest that the p.Lys254del mutation alone does not generate camptocormia or increase the CK level. However, future in-depth studies are required to unveil the underlying mechanisms related to the dominant pattern of inheritance and the phenotype of carriers of this *CAPN3* mutation.

## Data Availability

All data produced in the present study are available upon reasonable request to the authors

## ‘Declarations’

### Ethics approval and consent to participate

The study was approved by the Ethics Committee of the Hospital of origin; all subjects provided written informed consent.

### Consent for publication

All cases gave their consent.

### Availability of data and materials

The data that support the findings of this study are available from the corresponding author, upon reasonable request.

## Authors’ contributions

AS conceived the study. AV, GG-G, AM, CR-R, PC and AS carried out the experiments and the investigation. AV, GG-G, AM and AS performed the formal analysis. AV, GG, AM, CR-R, PC, ALdM and AS contributed in writing—review and editing the manuscript. GG, AM AS and ALdM supplied resources. All authors have read and agreed to the published version of the manuscript.

## Acknowledgements

The authors are grateful to the patients and the families.

## Abbreviations

LGMDR1: Limb-girdle muscular dystrophy recessive 1-calpain 3 related
LGMDD4: Limb-girdle muscular dystrophy dominant 4-calpain 3 related
MRI: Magnetic Resonance Image
OSAHS: Obstructive sleep apnoea hypopnoea syndrome.

